# Citation tracking for systematic literature searching: a scoping review

**DOI:** 10.1101/2022.09.29.22280494

**Authors:** Julian Hirt, Thomas Nordhausen, Christian Appenzeller-Herzog, Hannah Ewald

## Abstract

**Introduction:** Citation tracking (CT) collects references with citation relationships to pertinent references that are already known. This scoping review maps the benefit of and the tools and terminology used for CT in health-related systematic literature searching.

**Methods:** We included methodological studies on evidence retrieval by CT in health-related literature searching without restrictions on study design, language, or publication date. We searched MEDLINE/Ovid, Web of Science Core Collection, CINAHL/EBSCOhost, LLISFT/EBSCOhost, LISTA/EBSCOhost, conducted web searching via Google Scholar, backward/forward CT of included studies and pertinent reviews, and contacting of experts. Two reviewers independently assessed eligibility. Data extraction and analysis were performed by one reviewer and checked by another.

**Results:** We screened 11,861 references and included 47 studies published between 1985 and 2021. Most studies (96%) assessed the benefit of CT either as supplementary or stand-alone search method. Added value of CT for evidence retrieval was found by 96% of them. Science Citation Index and Social Sciences Citation Index were the most common citation indexes used. Application of multiple citation indexes in parallel, co-citing or co-cited references, CT iterations, or software tools was rare. CT terminology was heterogeneous and frequently ambiguous.

**Conclusions:** The use of CT showed an added value in most of the identified studies; however, the benefit of CT in health-related systematic literature searching likely depends on multiple factors that could not be assessed with certainty. Application, terminology, and reporting are heterogeneous. Based on our results, we plan a Delphi study to develop standard recommendations for the use and reporting of CT.

## Introduction

As systematic literature reviews aim at finding and synthesizing all available evidence on a topic,^1,2^ they are critical to inform health care practice and future research.^3-5^ Systematic reviews rely on information retrieval through systematic search strategies.^2^ It is challenging to design a systematic literature search that covers the ever-growing research volume, deals with the lack of universal terminology and indexation of research articles, and that keeps the number of results in an acceptable range for reviewers.^6-8^ According to current methodological guidance, systematic literature searching should apply both electronic database and supplementary search methods.^9^ In addition to contacting experts in the field, handsearching, trial registry searching, and web searching, supplementary methods include citation tracking (CT).^10^

CT exploits citation relationships to discover further eligible studies.^11^ While the methodological terminology around CT techniques is diverse,^12,13^ we will herein use CT as an umbrella term for multiple methods which collect related references from “seed references” through citation relationships.^11^ These seed references are references that are either known at the beginning of the review or emerge as eligible records following study selection and are usually eligible for inclusion in a review.^14,11^ CT methods can be sub-categorized into *direct* and *indirect* CT (for graphical representation see ^11^). For direct CT, the words “backward” and “forward” denote the directionality of tracking.^12,2^ Backward CT is the oldest form of CT. It identifies references that were cited by a seed reference which can be achieved at the title level by manually checking the reference list.^15,2^ In contrast, forward CT identifies citing references, i.e. references that cite a seed reference^11^ which can only be achieved by using an electronic citation index (e.g. Web of Science, Scopus, or Google Scholar). *Indirect* CT describes the identification of (i) co-cited references (i.e. publications sharing citing papers with a seed reference) and (ii) co-citing references (i.e. publications sharing references with a seed reference). Both direct and indirect CT may be conducted over one or more layers of iteration. To this end, researchers may use newly retrieved, relevant references as new seed references which we herein refer to as CT iterations.

The added value of any form of CT might not be the same for all systematic reviews. CT may be beneficial in research areas that require complex searches such as reviews of complex interventions, mixed-methods reviews, qualitative evidence syntheses, or reviews on public health topics. Research areas without consistent terminology or with vocabulary overlap with other fields, such as methodological topics, may also benefit from the use of CT.^10^ However, the use and benefit of CT in systematic literature searching as a basis for evidence-guided recommendations has not been systematically investigated.^11^ To fill this gap, we conducted this scoping review that was guided by the following three research questions:

- What is the benefit of citation tracking for systematic literature searching for health-related topics?
- Which methods, citation indexes, and tools are used for citation tracking?
- What terminology is used for citation tracking methods?

## Methods

A scoping review was conducted following the framework by Arksey and O’Malley^16,17^ and reported according to the “**P**referred **R**eporting **I**tems for **S**ystematic reviews and **M**eta-**A**nalyses extension for **Sc**oping **R**eviews” (PRISMA-ScR).^18^ A structured protocol has been published prospectively.^11^

### Eligibility criteria

We included any study that aimed at evaluating CT as an evidence retrieval method in a health-related context, if one of the following criteria was met: (i) Assessment of benefits/problems and/or effectiveness of CT, comparison of (ii) different methods of CT (e.g., backward vs. forward, direct vs. indirect) or (iii) technical uses of CT (e.g., citation indexes or tools). There were no restrictions on study design, language, and publication date.

We excluded studies that (i) solely applied but did not assess CT for evidence retrieval, (ii) assessed benefits and/or use and/or effectiveness of CT to explore a network or citation impact (i.e. bibliometric analyses), (iii) described the method of CT without further assessing it (e.g. guidelines for developing search strategies or for systematic or other review types), or (iv) only assessed the benefit of combined search methods in which the isolated benefit of CT could not be extracted. We also excluded editorials, commentaries, letters, and abstract-only publications. Any type of literature review was included to search for primary studies but excluded from our analysis.

### Information Sources

We searched MEDLINE via Ovid, CINAHL (Cumulative Index to Nursing and Allied Health Literature), LLISFT (Library Literature & Information Science Full Text) and LISTA (Library, Information Science & Technology Abstracts) via EBSCOhost, and the Web of Science Core Collection on October 26, 2020 (see supporting information 1). As supplementary search methods, we performed web searching via Google Scholar (on December 7, 2020) using search terms from our database search as well as direct forward and backward CT of included studies and pertinent review articles that were flagged during the screening of search results (on February 10, 2021). For forward CT, we used Scopus, Web of Science, and Google Scholar. For backward CT, we used Scopus and, if seed references were not indexed in Scopus, we manually extracted the seed references’ reference list. We iteratively repeated forward and backward CT on newly identified eligible references until no further eligible references or pertinent reviews could be identified (three iterations; the last iteration on May 5, 2021). We also contacted librarians in the field of health sciences and information specialists through four mailing lists (Canadian Medical Libraries, Expertsearching, MEDIBIB-L/German-speaking medical librarians, and EAHIL-list) for further eligible studies.

### Search Strategy

HE drafted the search strategies and JH checked them according to the Peer-Review of Electronic Search Strategies (PRESS) guideline.^19^ We limited the strategy to text words due to a lack of adequate index terms. To determine frequently occurring terms for inclusion in the search strategy, we analyzed keywords in the titles and abstracts of potentially relevant publications retrieved from preliminary searches and similar articles identified via PubMed by using various text mining tools (PubMed Reminer, AntConc, Yale MeSH analyzer, Voyant, VOSviewer, Termine, Text analyzer).^20^ We restricted some text words to the title field in order to avoid retrieving systematic reviews that used CT. Translation of the original PubMed strategy to other syntax was done using the Polyglot Search Translator.^21^ For topical restriction and reasons of feasibility, the retrieval from Web of Science was limited to pertinent Web of Science Categories and Research Areas. Final search strategies are reported in supporting information 1.

### Data management

CAH conducted the database searches and CT, exported results to EndNote X9 (Clarivate), and eliminated duplicates using the Bramer method.^22^ JH conducted the web search and contacted the experts. Citavi was used to manage the number of reference retrievals throughout the study selection process.^23^ Additionally, we used specific tools for study selection that we describe below.

### Selection of sources of evidence

After an initial calibration phase of screening 100 titles and abstracts separately and discussing divergent decisions (TN, JH, HE), two authors (JH, TN) independently screened titles, abstracts, and full texts using the web-app Rayyan.^24^ Disagreements were solved by third author arbitration (HE or CAH).

### Data charting process

We used a prespecified data extraction spreadsheet (ONLYOFFICE, SWITCHdrive^25^) that was approved by consensus among the authors. We extracted bibliographic and geographic data (reference, publication year, and affiliated countries), design- and study-specific data (purpose of CT, health context, test sample, CT methods [e.g., backward CT, forward CT], terminology to describe CT, CT iterations, reported citation indexes, reported CT tools, outcome comparison[s] and measure[s]) as well as study results and authors’ conclusions. One author (JH, TN, CAH, or HE) extracted data and a second author (JH, TN, CAH, or HE) peer-checked the extraction. We solved disagreements by third author arbitration (one out of JH, TN, CAH, or HE).

### Synthesis of results

One author (JH, TN, CAH, or HE) narratively summarized and tabulated the study characteristics and results using numbers and percentages based on the data extraction. We aimed to provide a synthesis of any benefit of using CT. For operationalization, we checked authors’ discussions and conclusions for clear statements of an added value or no added value. If clear statements were not present, we examined the study results: In studies where CT was used as a supplementary search method, we scored an added value if the use of CT compared to another search technique retrieved unique references. In studies where CT was used as a stand-alone search method, we scored an added value if CT identified more eligible references than the comparator search method or reduced the screening load. We additionally analyzed if an added value was brought into a specific context by the authors.

## Results

### Characteristics of included studies

Database and supplementary searches yielded 11,861 unique references. After title-abstract screening, we assessed 221 references in full text. Of these, we excluded 171 references due to various reasons, mostly wrong study aim, and finally included 47 studies^26-72^ (Figure 1). For three of the studies,^41,71,34^ we found related documents (erratum,^73^ doctoral thesis,^74^ and evidence summary^75^), yielding a total of 50 reports, two of which were conference posters.^50,45^ Twenty-six reports (52%) were included from the results of the bibliographic database search and 24 (48%) from the supplementary search results (one from web searching, 14 from backward CT, six from forward CT, and three from contacting experts). Further CT iterations did not yield any additional records that met our inclusion criteria.

**Figure 1.**
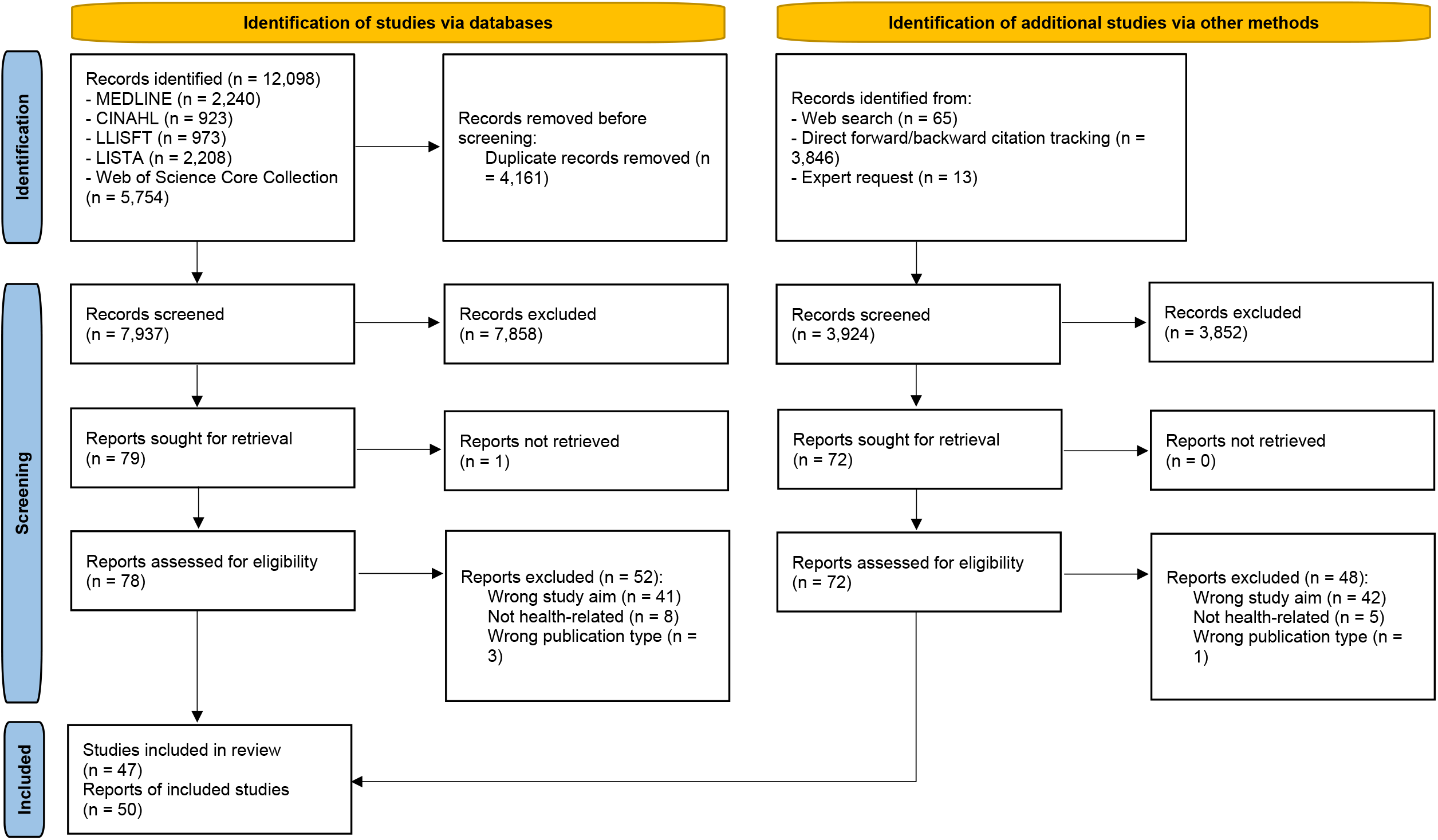
Literature search and study retrieval process (PRISMA 2020 flow diagram) Abbreviations: CINAHL = Cumulative Index to Nursing and Allied Health Literature; LISTA = Library, Information Science and Technology Abstracts; LLISFT = Library Literature & Information Science Full Text; MEDLINE = Medical Literature Analysis and Retrieval System Online.

The 47 studies were published between 1985 and 2021; the median publication year was 2014. Most of the studies were nationally authored without international collaboration. More than 70% (34 studies) of the studies were conducted by authors from the UK or the US. Most units of analysis in the included studies (in 68% of studies one or more systematic reviews) dealt with a single medical field or health topic (35 studies, 74%) rather than various fields or topics (nine studies, 19%). Twenty-seven studies (57%) dealt with research questions that assessed interventions (Table 1).

**Table 1.** Basic characteristics of the included studies (n=47)

|  |  | n (%) |
| --- | --- | --- |
| Publication decade | 1980s | 5 (11) 52,58,61,62,70 |
|  | 1990s | 5 (11) 43,51,53,59,60 |
|  | 2000s | 10 (21) 26,36,44,46,47,54,55,57,67,68 |
|  | 2010s | 19 (40) 27,29,30,32-34,38,41,45,48-50,56,64,66,69,71,72,63 |
|  | 2020s * | 8 (17) 35,28,31,37,39,40,42,65 |
| Authorship ** | Internationally authored | 4 (9) 32,35,39,56 |
|  | Nationally authored | 43 (92) 26-31,33,34,36-38,40-49,51,50,52,54,53,55,57-72 |
| Affiliated countries ** | UK | 17 (36) 26,36,51,53,54,57,63-65,68,70,72,34,48,44,50,38 |
|  | US | 17 (36) 32,37,47,55,58-62,29,30,40-42,49,52,66 |
|  | Germany | 5 (11) 56,45,71,35,39 |
|  | Canada | 3 (6) 35,31,67 |
|  | Austria | 2 (4) 32,56 |
|  | Netherlands | 2 (4) 28,43 |
|  | Switzerland | 2 (4) 39,56 |
|  | Australia | 1 (2) 33 |
|  | Denmark | 1 (2) 46 |
|  | Korea | 1 (2) 27 |
|  | Romania | 1 (2) 56 |
|  | Tunisia | 1 (2) 69 |
| Unit of analysis in included studies (with number included in each) | Systematic reviews (n=1 to n=250 ***) | 32 (68) 27,36,45,68,37,55,63,57,26,54,71,53,51,31,32,34,50,40,49,48,38,44,28-30,67,43,42,66,64,56,41 |
|  | Systematic and non-systematic reviews (n=20) | 1 (2) 33 |
|  | Overviews of reviews (n=86) | 1 (2) 35 |
|  | Scoping review (n=1) | 3 (6) 39,72,65 |
|  | Search strategies (n=1 to n=89) | 6 (13) 46,47,69,59,60,52 |
|  | Set of references (n=111 to n=1331) | 4 (9) 61,62,58,70 |
| Medical field/health topic of sample studies | Mental or cognitive health/psychology | 9 (19) 71,37,45,39,70,72,40,34,65 |
|  | Orthopedics | 6 (13) 55,69,31,57,46,26 |
|  | Gastroenterology | 3 (6) 32,58,62 |
|  | Hematology/Oncology | 3 (6) 27,68,47 |
|  | Cardiology/Angiology | 2 (4) 43,44 |
|  | Endocrinology | 1 (2) 50 |
|  | Infectiology | 1 (2) 33 |
|  | Pulmonology | 1 (2) 61 |
|  | Health decision/policy making | 4 (9) 49,54,38,36 |
|  | Health diagnostics | 3 (6) 53,48,64 |
|  | Health communication/education | 2 (4) 63,51 |
|  | Various medical fields/health topics | 9 (19) 29,30,66,67,35,41,42,56,52 |
|  | Not reported | 3 (6) 59,60,28 |
| Intervention characteristic of sample studies **** | Clinical intervention | 21 (45) 43,50,58,62,32,68,51,49,53,64,33,71,37,45,55,69,26,29,30,66,67 |
|  | Non-clinical intervention | 6 (13) 63,36,34,65,72,57 |
|  | No intervention | 10 (21) 44,27,47,38,54,48,39,70,40,31 |
|  | Various | 6 (13) 46,35,41,42,56,52 |
|  | Not reported | 4 (9) 28,59,61 |

Abbreviations: UK = United Kingdom; US = United States (of America).
\* Note that database and supplementary searches were performed between October 2020 and May 2021.
\*\* Considering all affiliated authors and countries.
\*\*\* Number per included study is reported in the raw data, see supporting information 2.
\*\*\*\* Clinical intervention: Health-related measures on a patient/population-level (e.g., diagnosis, therapy, care, prevention) focusing on specific diseases.
Non-clinical intervention: Health-related measures on organizational or health policy and research level, health-related measures on patient/population-level not focusing on specific diseases (e.g., common health behavior).
No intervention: Studies that do not assess interventions (e.g., epidemiological, prognostic, methodological or descriptive studies).
Various: Mix of at least two of the three characteristics "clinical intervention", "non-clinical intervention", and "no intervention".
Not reported: No information provided in the eligible study.

### Benefit of citation tracking

Forty-five studies (96%) assessed the benefit of CT. Of these, 25 studies (56%) performed supplementary CT following prior database searching and 20 studies (44%) stand-alone CT, including four studies that used stand-alone CT for a review update.^40,50,27,44^ Mostly, the performance of CT was compared to a search in multiple databases (21 studies, 47%). Typical outcome measures were the number of retrieved eligible articles (27 studies, 60%), unique articles that were only identified by CT (8 studies, 18%), and/or retrieved articles (7 studies, 16%). A benefit of CT was usually ascribed by the authors if the results of CT significantly contributed to these outcome measures or if methodological efficiency of evidence retrieval (i. e. the relevancy and precision of the output) was increased (Table 2 and supporting information 2). Notably, while only two (4%) studies that assessed the benefit of CT found no added value of the use of CT,^71,72^ 40% of those authors that stated an added value brought it into a specific context. Thus, particular study designs (observational, prognostic, or diagnostic test studies) or research topics such as non-pharmacological, non-clinical, public health, or alternative medicine, which may be regarded as complex, broad, fringe, or difficult-to-locate, were proposed to benefit most from CT (Table 2).

**Table 2.** Study focus of included studies (n=47) and purpose of CT, comparator, outcome measure, and added value of CT in studies assessing the benefit of CT (n=45)

|  |  | n (%) |
| --- | --- | --- |
| Study focus (n=47) * | Benefit of CT | 45 (96) 26,28-32,34-47,49-72,27 |
|  | Technical uses of CT | 8 (17) 33,32,27,28,65,72,48,49 |
|  | Different methods of CT | 5 (11) 45,27,36,63,29 |
| Purpose of CT in relation to database search (n=45) | Supplementary search | 25 (56) 26,31,35-37,39,43,45-47,51,53,55-57,59,63-65,67,68,70-72,54 |
|  | Primary/stand-alone search | 20 (44) 58,60,61,44,28-30,32,38,40-42,49,50,52,66,69,62,34,27 |
| Comparator (n=45) | Multiple database search | 21 (47) 35-37,39,40,45,47,49,50,52,55-58,61-64,68,70,72 |
|  | Multiple database search and supplementary search methods ** | 5 (11) 26,51,53,67,71 |
|  | Single database search | 9 (20) 31,38,43,44,46,59,60,69,27 |
|  | Supplementary search methods | 1 (2) 54 |
|  | Multiple search methods *** | 9 (20) 54,29,30,28,34,32,41,42,65,66 |
| Outcome measure (n=45) * | Number of retrieved eligible articles | 27 (60) 58,62,28,46,35,45,52,57,63,64,70,26,53,51,71,41,66,54,59,43,55,68,36,50,32,38,27 |
|  | Number of unique articles | 8 (18) 37,39,34,44,31,65,72,47 |
|  | Number of retrieved articles | 7 (16) 59,61,40,31,40,42,59,67,28,61 |
|  | Other **** | 6 (13) 56,29,49,60,30,69 |
| Added value of CT (n=45) | Yes | 43 (96) 26,28-32,34-47,49-70,27 |
|  | No | 2 (4) 71,72 |
| If added value, added value brought into specific context by the authors (n=17) | Diagnostic test studies | 2 (4) 50,64 |
|  | Observational studies | 2 (4) 47,44 |
|  | Complex research questions ***** | 2 (4) 36,45 |
|  | Health services and public health | 2 (4) 54,57 |
|  | Depending on the topic | 1 (2) 26 |
|  | Disciplines and topics in which citations are numerous | 1 (2) 29 |
|  | Broader scope | 1 (2) 59 |
|  | Fringe areas | 1 (2) 61 |
|  | Non-published and difficult to locate studies ***** | 1 (2) 68 |
|  | Prognostic studies | 1 (2) 31 |
|  | Non-clinical research | 1 (2) 70 |
|  | Non-pharmacological interventions | 1 (2) 56 |
|  | Alternative medicine | 1 (2) 43 |

Abbreviations: CT = Citation tracking.
\* More than one category possible.
\*\* E.g., hand-searching, contacting experts.
\*\*\* Potentially multiple search methods used which were not described in detail, e.g., “Cochrane review methods” without providing details.
\*\*\*\* E.g., time, relevancy, precision (see supporting information 2 for details).
\*\*\*\*\* E.g., as those undertaken for management and policymaking questions.
\*\*\*\*\* E.g., conference abstracts, technical reports.

### Methods, citation indexes, and tools used for citation tracking

With respect to CT methods, 33 studies (70%) assessed backward CT, whereas forward CT was somewhat less frequently assessed (29 studies, 62%). Indirect CT methods were assessed in 7/47 studies (15%) for co-cited and 6/47 studies (13%) for co-citing CT (Table 3).

**Table 3.**
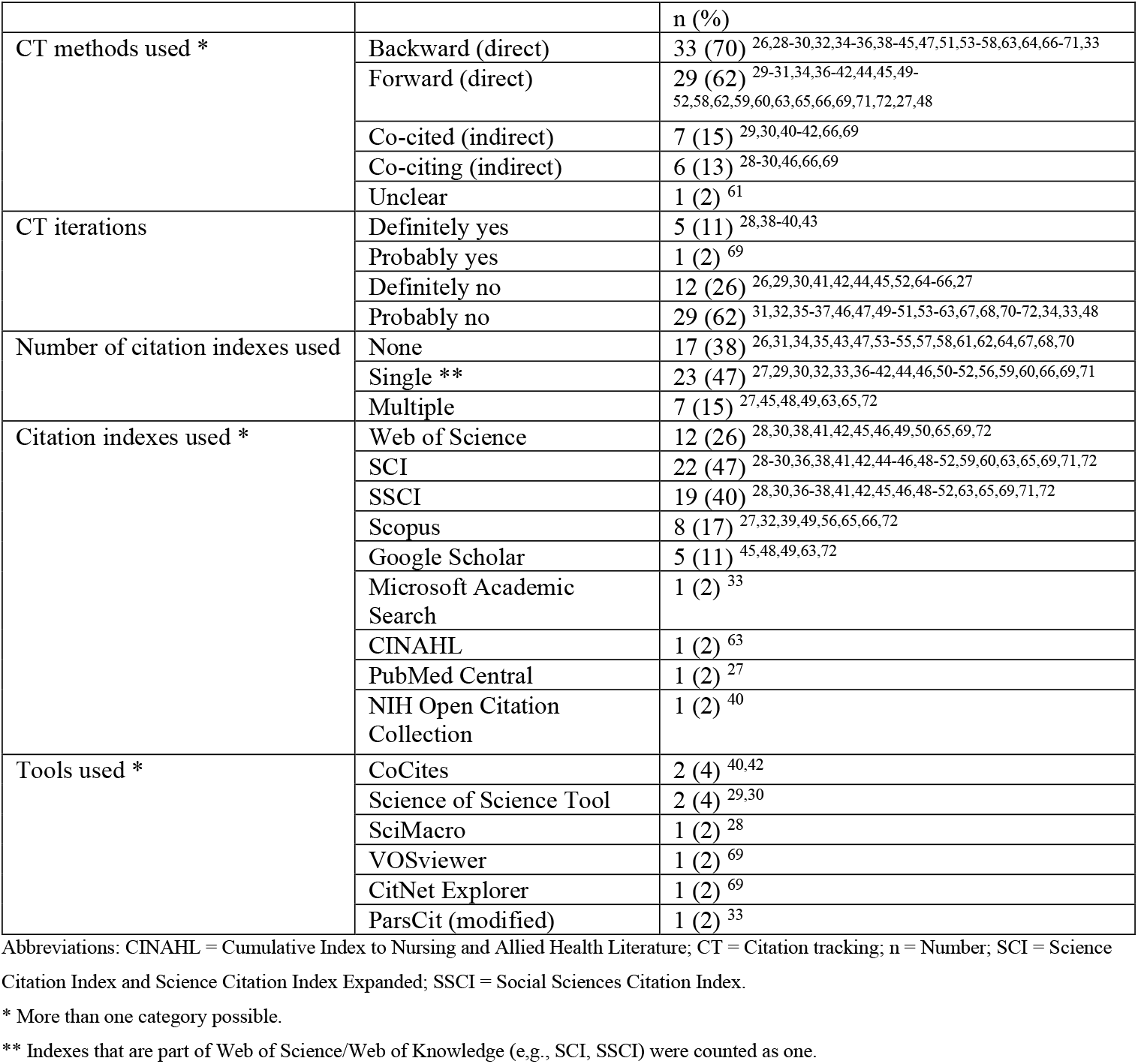
Methodological characteristics of the included studies (n=47)

Eight studies reported comparisons of different CT methods.^36,44,45,51,63,29,27,71^ In three studies, backward CT retrieved more eligible references than forward CT.^36,51,45^ In two studies, forward CT retrieved more eligible references than backward CT^44,63^ (supporting information 2). The relative performance of indirect CT methods is reported in a separate paragraph (see below).

Although the associated reporting was unclear, most studies definitely (twelve studies, 26%) or at least probably (29 studies, 62%) omitted CT iterations and performed only one round of CT (Table 3).

Seventeen studies (36%) performed CT (mainly backward) without the use of a citation index. Of the 30 studies that used at least one citation index, only seven (23%) used and compared different citation indexes. By far the most popular citation indexes were Science Citation Index/Science Citation Index Expanded (SCI, 22 studies) and Social Sciences Citation Index (SSCI, 19 studies) that were used by themselves or in context of the Web of Science Core Collection (twelve studies). Other citation indexes included Scopus (eight studies) and Google Scholar (five studies) (Table 3). Authors that compared CT indexes found that, compared to Scopus or Web of Science, the use of Google Scholar for forward CT is associated with a high administrative and time cost,^48,72^ which led other authors to exclude Google Scholar results “for practical reasons”.^45^ In terms of yield of forward CT, Scopus was reported to be superior to Web of Science.^65^ Likewise, the forward CT results from Scopus were more precise than those from Web of Science or Google Scholar when searching for a specific diagnostic test^49^ (supporting information 2).

Several studies designed or applied software tools for CT (Table 3).

### Indirect citation tracking

Relatively little evidence exists regarding the utility of indirect CT for health-related evidence retrieval. Nonetheless, the replication of fourteen Cochrane reviews by combined CT methods suggested that co-cited references may offer better coverage of relevant literature compared to cited, citing, or co-citing references.^29^ Independent work, which led to the development of the CT software CoCites,^42^ also documented the effectiveness of co-cited references for health-related evidence retrieval.^41^ Moreover, the studies on indirect CT revealed and pioneered the necessity for various relevancy ranking methods to prioritize and reduce the abundant output of indirect CT (supporting information 2).^41,66,29,30^

### Terminology used for citation tracking methods

We extracted the terminology that was used by study authors for CT methods and for seed references. As documented in Table 4, terminology was heterogeneous and, in some instances, ambiguous.

**Table 4.**
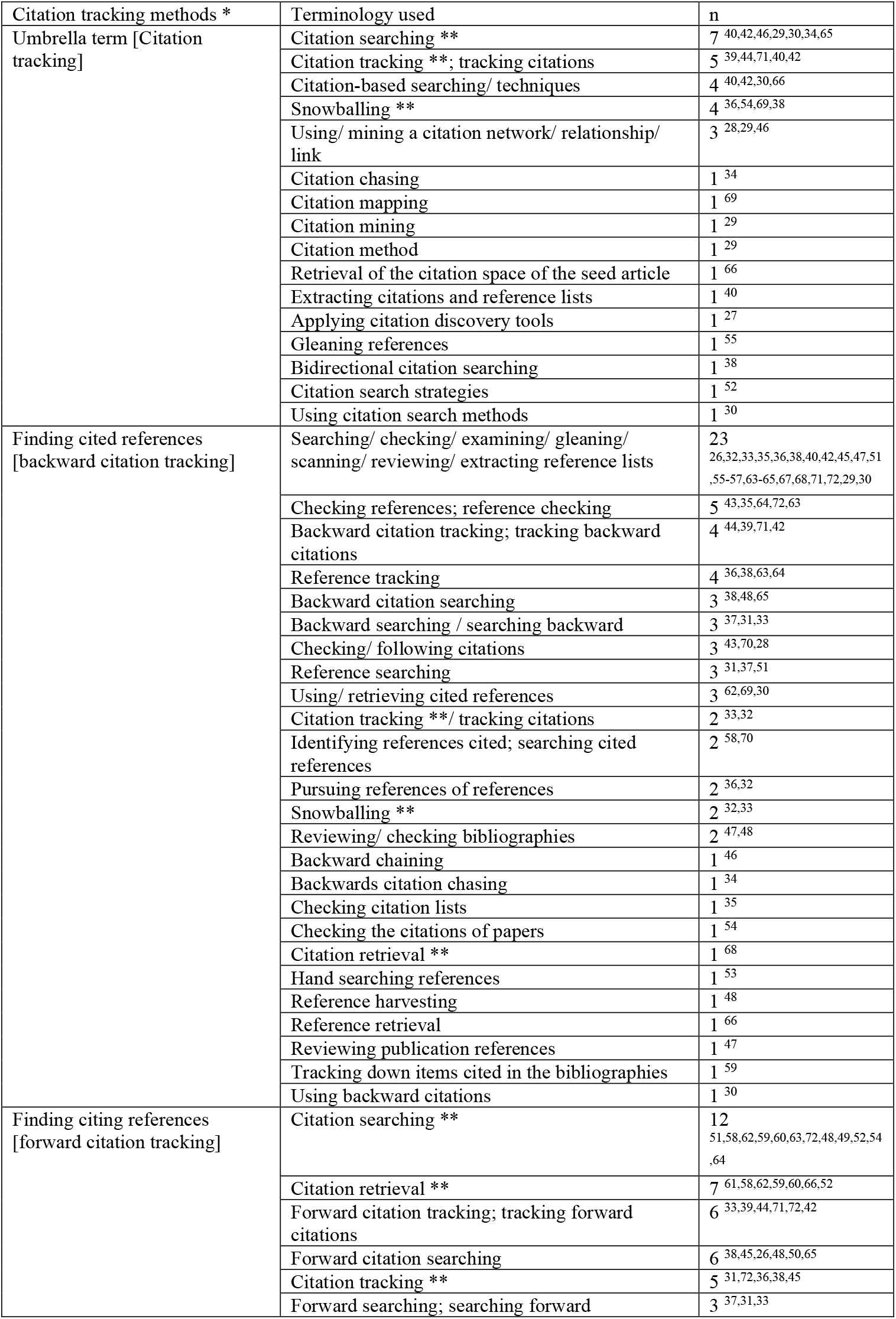
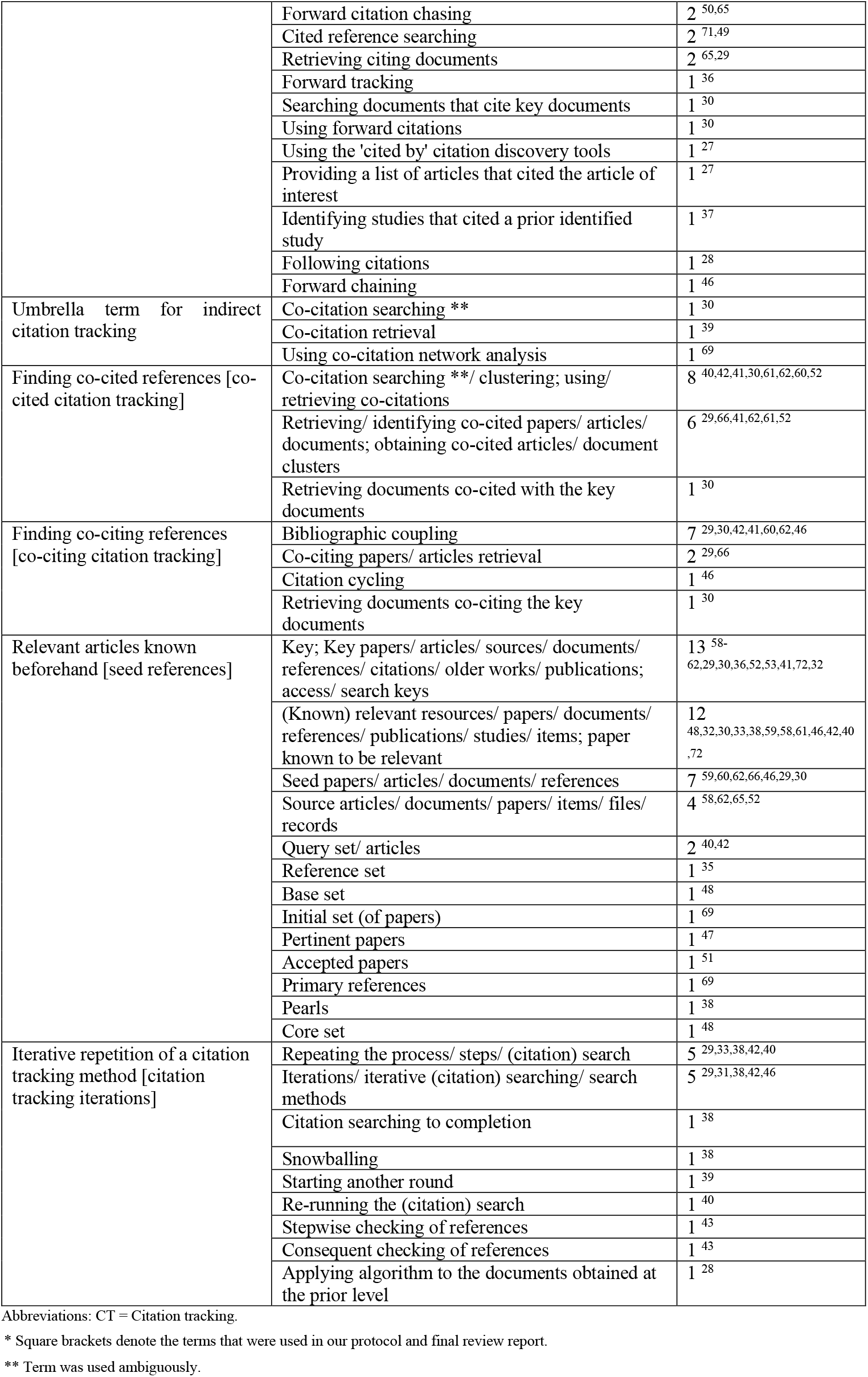
Terminology used for citation tracking methods, ordered by frequency

| Citation tracking methods * | Terminology used | n |
| --- | --- | --- |
| Umbrella term [Citation tracking] | Citation searching ** | 7 40,42,46,29,30,34,65 |
|  | Citation tracking **: tracking citations | 5 39,44,71,40,42 |
|  | Citation-based searching/ techniques | 4 40,42,30,66 |
|  | Snowballing ** | 4 36,54,69,38 |
|  | Using/ mining a citation network/ relationship/ link | 3 28,29,46 |
|  | Citation chasing | 1 34 |
|  | Citation mapping | 1 69 |
|  | Citation mining | 1 29 |
|  | Citation method | 1 29 |
|  | Retrieval of the citation space of the seed article | 1 66 |
|  | Extracting citations and reference lists | 1 40 |
|  | Applying citation discovery tools | 1 27 |
|  | Gleaning references | 1 55 |
|  | Bidirectional citation searching | 1 38 |
|  | Citation search strategies | 1 52 |
|  | Using citation search methods | 1 30 |
| Finding cited references [backward citation tracking] | Searching/ checking/ examining/ gleaning/ scanning/ reviewing/ extracting reference lists | 23 26,32,33,35,36,38,40,42,45,47,51,55-57,63-65,67,68,71,72,29,30 |
|  | Checking references; reference checking | 5 43,35,64,72,63 |
|  | Backward citation tracking; tracking backward citations | 4 44,39,71,42 |
|  | Reference tracking | 4 36,38,63,64 |
|  | Backward citation searching | 3 38,48,65 |
|  | Backward searching / searching backward | 3 37,31,33 |
|  | Checking/ following citations | 3 43,70,28 |
|  | Reference searching | 3 31,37,51 |
|  | Using/ retrieving cited references | 3 62,69,30 |
|  | Citation tracking **: tracking citations | 2 33,32 |
|  | Identifying references cited; searching cited references | 2 58,70 |
|  | Pursuing references of references | 2 36,32 |
|  | Snowballing ** | 2 32,33 |
|  | Reviewing/ checking bibliographies | 2 47,48 |
|  | Backward chaining | 1 46 |
|  | Backwards citation chasing | 1 34 |
|  | Checking citation lists | 1 35 |
|  | Checking the citations of papers | 1 54 |
|  | Citation retrieval ** | 1 68 |
|  | Hand searching references | 1 53 |
|  | Reference harvesting | 1 48 |
|  | Reference retrieval | 1 66 |
|  | Reviewing publication references | 1 47 |
|  | Tracking down items cited in the bibliographies | 1 59 |
|  | Using backward citations | 1 30 |
| Finding citing references [forward citation tracking] | Citation searching ** | 12 51,58,62,59,60,63,72,48,49,52,54,64 |
|  | Citation retrieval ** | 7 61,58,62,59,60,66,52 |
|  | Forward citation tracking; tracking forward citations | 6 33,39,44,71,72,42 |
|  | Forward citation searching | 6 38,45,26,48,50,65 |
|  | Citation tracking ** | 5 31,72,36,38,45 |
|  | Forward searching; searching forward | 3 37,31,33 |
|  | Forward citation chasing | 2<sup>50,65</sup> |
|  | Cited reference searching | 2<sup>71,49</sup> |
|  | Retrieving citing documents | 2<sup>65,29</sup> |
|  | Forward tracking | 1<sup>36</sup> |
|  | Searching documents that cite key documents | 1<sup>30</sup> |
|  | Using forward citations | 1<sup>30</sup> |
|  | Using the 'cited by' citation discovery tools | 1<sup>27</sup> |
|  | Providing a list of articles that cited the article of interest | 1<sup>27</sup> |
|  | Identifying studies that cited a prior identified study | 1<sup>37</sup> |
|  | Following citations | 1<sup>28</sup> |
|  | Forward chaining | 1<sup>46</sup> |
| Umbrella term for indirect citation tracking | Co-citation searching ** | 1<sup>30</sup> |
|  | Co-citation retrieval | 1<sup>39</sup> |
|  | Using co-citation network analysis | 1<sup>69</sup> |
| Finding co-cited references [co-cited citation tracking] | Co-citation searching **/ clustering; using/ retrieving co-citations | 8<sup>40,42,41,30,61,62,60,52</sup> |
|  | Retrieving/ identifying co-cited papers/ articles/ documents; obtaining co-cited articles/ document clusters | 6<sup>29,66,41,62,61,52</sup> |
|  | Retrieving documents co-cited with the key documents | 1<sup>30</sup> |
| Finding co-citing references [co-citing citation tracking] | Bibliographic coupling | 7<sup>29,30,42,41,60,62,46</sup> |
|  | Co-citing papers/ articles retrieval | 2<sup>29,66</sup> |
|  | Citation cycling | 1<sup>46</sup> |
|  | Retrieving documents co-citing the key documents | 1<sup>30</sup> |
| Relevant articles known beforehand [seed references] | Key; Key papers/ articles/ sources/ documents/ references/ citations/ older works/ publications; access/ search keys | 13<sup>58-62,29,30,36,52,53,41,72,32</sup> |
|  | (Known) relevant resources/ papers/ documents/ references/ publications/ studies/ items; paper known to be relevant | 12<sup>48,32,30,33,38,59,58,61,46,42,40,72</sup> |
|  | Seed papers/ articles/ documents/ references | 7<sup>59,60,62,66,46,29,30</sup> |
|  | Source articles/ documents/ papers/ items/ files/ records | 4<sup>58,62,65,52</sup> |
|  | Query set/ articles | 2<sup>40,42</sup> |
|  | Reference set | 1<sup>35</sup> |
|  | Base set | 1<sup>48</sup> |
|  | Initial set (of papers) | 1<sup>69</sup> |
|  | Pertinent papers | 1<sup>47</sup> |
|  | Accepted papers | 1<sup>51</sup> |
|  | Primary references | 1<sup>69</sup> |
|  | Pearls | 1<sup>38</sup> |
|  | Core set | 1<sup>48</sup> |
| Iterative repetition of a citation tracking method [citation tracking iterations] | Repeating the process/ steps/ (citation) search | 5<sup>29,33,38,42,40</sup> |
|  | Iterations/ iterative (citation) searching/ search methods | 5<sup>29,31,38,42,46</sup> |
|  | Citation searching to completion | 1<sup>38</sup> |
|  | Snowballing | 1<sup>38</sup> |
|  | Starting another round | 1<sup>39</sup> |
|  | Re-running the (citation) search | 1<sup>40</sup> |
|  | Stepwise checking of references | 1<sup>43</sup> |
|  | Consequent checking of references | 1<sup>43</sup> |
|  | Applying algorithm to the documents obtained at the prior level | 1<sup>28</sup> |

Abbreviations: CT = Citation tracking.
\* Square brackets denote the terms that were used in our protocol and final review report.
\*\* Term was used ambiguously.

## Discussion

By using up-to-date methodology of systematic evidence retrieval and synthesis, we provide the first comprehensive analysis of the use and benefit of CT in systematic literature searching. Focusing on health-related literature, we identified 47 methodological studies. CT is a research method that has been used for systematic literature searching for almost 40 years. Nevertheless, detailed methodological guidance for its use and reporting in evidence retrieval is largely missing. The present scoping review which maps the current evidence on CT is the first milestone in a larger research endeavor to develop such guidance.^11^

Forty-three of the 45 studies that assessed the benefit of CT found added value of CT for evidence retrieval. It is tempting to conclude from this that CT is of paramount importance for systematic evidence synthesis in health-related topics. However, these numbers should be interpreted with caution: First, included studies displayed highly heterogeneous methodological and topical characteristics that could influence the benefit of CT (e.g., unit of analysis in included studies, quality of comparator search method, health topic, CT methods used, number of CT iterations, or citation indexes and tools used). Second, we believe that the number 2 versus 43 in fact underestimates the ratio of no added value versus added value of CT due to publication bias. Thus, researchers who applied CT but did not find an added value may be significantly less likely to publish their results in a methodology paper.

Based on these considerations, it is important to highlight the specific features and conclusions of the two studies that reported no added value of CT.^71,72^ Westphal et al. conducted a systematic review of randomized controlled trials on the efficacy of psychotherapeutic, pharmacological, and combined interventions in the treatment of chronic depression. They searched seven bibliographic databases and identified 2417 unique records. The authors also performed a variety of supplementary search methods yielding >27,000 records. They concluded that hand-searching contents of relevant journals and screening reference lists of related systematic reviews were effective but backward and forward CT on included records using SCI and SSCI was not because it did not lead to any further inclusion of primary studies.^71^ Wright et al. performed six sensitive database searches yielding almost 22,000 records on interventions targeting change in at least two risk behaviors. Their scoping review searches were complemented with laborious forward CT on the 40 included papers in Google Scholar, Scopus, Web of Science, and Ovid MEDLINE. This elaborate CT search found only one eligible paper that was not previously identified by database searching. The authors concluded that “citation searching as a supplementary search method for systematic reviews may not be the best use of valuable time and resources”.^72^

While it would be desirable for researchers to know exactly in which situations a possible added value of CT would or would not outweigh the increased workload that comes with it, a clean categorization is currently not possible. But analyzing the contexts of the eligible CT studies, we may have found other factors that could play a role. For instance, CT may be less beneficial in situations where researchers operate with clearly defined clinical interventions as part of Participant-Intervention-Comparison-Outcome (PICO)-questions than with hard-to-search-for topics such as non-clinical intervention or policymaking questions.^36^ Concerning potential correlations between research topic and added value of CT, we realized that “CT for systematic literature searching” was a hard-to-search-for topic when we composed the search strings for this scoping review. Thus, sensitive versions of database search strings would have returned far too many results, necessitating pragmatic search decisions (see section limitations). Consistent with the above observations, the CT approaches applied as supplementary searches resulted in the identification of 40% of our included studies and critically added value to our work. But taking a bird’s eye view and as outlined by Horsley et al.,^76^ it is currently challenging for reviewers to recognize situations when database searches are not sufficiently exhaustive and should be supplemented by CT methods.

CT was conducted as a primary or stand-alone search approach instead of a supplementary search method in almost half of the methodological studies collected in this scoping review. For this purpose, searchers would retrieve a handful of seed references from private collections or simplified database searches and use them as a basis for stand-alone CT.^69^ Reviewing all the studies comparing the effectiveness of stand-alone CT with database searching reveals that the former method rarely finds as many relevant articles as the comparator search strategy.^29,42,41,38,44,49,50,52,61,66^ This leads us to the conclusion that stand-alone CT appears not to be sensitive enough for systematic reviews and scoping reviews or their updates. Having said that, optimized stand-alone CT methods may prove an interesting alternative to database searching for narrative, rapid, or systematized reviews and for researchers composing research-in-context assessments or grant applications. For complex topics that are unamenable to subject searches (see above), stand-alone CT techniques could also be considered as a pragmatic workaround search approach.

Concerning different CT methods, we found that almost five times as many methodological studies assessed direct as indirect CT. Out of direct CT assessments, about as many studies as have assessed forward have also examined backward CT. Since backward CT is used far more frequently than forward CT,^15^ we were surprised that there were not more methodology papers assessing it. It is possible that reviewers who apply backward CT, the oldest CT method, usually do not analyze the results of it in a separate methodology paper. Regarding indirect CT, the few included studies that applied and assessed indirect CT methods strongly hinted its potential utility for health-related evidence retrieval.^29,41,42^ However, as these methods usually lead to an abundant output, ranking methods to prioritize this output according to relevance are necessary.^41,66,29,30^ Facing only a few studies that evaluated indirect CT as stand-alone methods in our sample, we propose that the development of recommendations for applicability and conduct of indirect CT methods as supplementary search methods as well as ranking methods for reference retrieval requires further research.

SCI and SSCI were the most common citation indexes used to conduct CT. Application of multiple citation indexes in parallel was rare. However, all studies that applied more than one citation index found that the results of the different indexes complemented each other.^72,48,65^ Collecting CT results systematically from several citation indexes therefore enhanced the coverage of citations which is somewhat reminiscent of the complementary effect of using multiple databases for searching. This applies to both forward and backward CT. Backward CT is still frequently being performed manually by screening reference lists. However, performing backward CT using electronic citation indexes in combination with reference management software is preferable, since it allows deduplication of the references against each other and against the results of the primary search as well as effective title and abstract screening.^32,77^

Only a minority of systematic reviewers perform CT iterations. Three of the included methodological studies that performed CT iterations reported unique relevant publications that were identified only during the second or third CT iteration.^38,43,40^ In our scoping review, we performed three CT iterations. The first iteration yielded twenty papers. Although the second iteration, based on those did not yield any new eligible references, it did identify a SuReInfo chapter^78^ and a precursor paper of one of our includes^52^ that so far escaped our searches and were used as seed references for a third CT iteration. The third iteration yielded no more relevant papers. Thus, there is evidence that conducting CT iterations can contribute to the comprehensiveness of a systematic search.

Only a few identified methodological studies reported specific software tools for CT automation. While CT automation could be more time-saving and practical in general, detailed assessments would be needed to measure time-savings, recall or potential costs. During the finalization of this scoping review, two new and publicly available tools have been published: the *citationchaser*^79^ for automated forward and backward CT and the *Citation Cloud* PubMed extension^80^ for automated forward, backward, co-cited, and co-citing CT.

On a more general note, we found that the reporting of CT methods is frequently unclear and far from being standardized. A possible reason for this could be the lack of specific guidance for the conduct and reporting of CT as current gold-standard guidelines for systematic reviews are relatively lax as far as CT is concerned.^2,15,81^ High heterogeneity is also reflected by the obvious nonuniformity of CT terminology. Several terms are used ambiguously and it is often unclear what they stand for. E.g., “citation searching”, “snowballing”, or “co-citation searching” are sometimes used for the methodological umbrella term but also for a specific method such as backward or forward CT. Furthermore, the use of CT is not restricted to systematic literature searching in health research. There are other fields where CT methods are applied, e.g., bibliometric research which explores citation networks based on authors, institutions, countries, or topics.^46,81-83^ This may partly explain the existence of various terms that can be used for CT methods.

Taken together, a rich albeit heterogeneous evidence landscape exists regarding the use of CT in health-related systematic literature searching which spans decades of common practice. The present scoping review is a first attempt to systematically synthesize this evidence. Our results make a strong case for the urgent need for evidence-based and researcher-approved guidelines for the use of CT.

### Limitations

Our scoping review has several limitations. First, we did not consider articles that were at preprint status at the time of study selection^84^ which would have led to the inclusion of further studies.^85^ Second, during the work on this review, we became aware of “bibliographic coupling” as a relevant term that was missing from our search strategy, which possibly led to the omission of eligible articles and should be reconsidered for updates of this review. Third, our decision to limit the Web of Science search to pertinent Web of Science Categories and Research Areas was pragmatic and potentially incompatible with systematic retrieval. Fourth, we did not assess the quality and sensitivity of the database searches in included studies. This could be considered in future studies since the quality and sensitivity of database searching as a primary or comparator search method may indirectly influence the effectiveness of CT. Fifth, the dichotomous way we scored “added value yes/no” from heterogeneous data as a composite outcome of author statements and our own definition of added value neither reflected the size of that value (e.g., how many more (unique) eligible references does CT find than the comparator?) nor its usefulness (e.g., does finding these extra studies change the results of meta-analysis?). Sixth, we restricted eligibility to methodological studies with a focus on CT as an evidence retrieval method. This almost certainly led to the neglect of (systematic) reviews with an implicit evaluation of the benefit of CT, e.g., as indicated by the detailed documentation of article retrieval sources.^86^ Seventh, as our scoping review eligibility was restricted to health sciences, we neglected the methodological studies of other fields that assessed CT. While the benefits of CT could differ between fields (as it likely does between topics), our main reason for this restriction was feasibility, i.e., to reduce the massive amounts of search results. This seemed especially serious for software tools for CT that were developed with an interdisciplinary scope. Hence, the list of identified methodological studies and software tools is clearly not exhaustive.^87,88^ Finally, we did not request information about the isolated results of CT from authors who applied and evaluated several supplementary search methods together. This might have led to the inclusion of a few further studies^89-91^ and should be considered for updates of this review.

## Conclusions

Our scoping review features a broad body of studies investigating the use of CT as a literature search method for health-related topics. We found large heterogeneity regarding its application, terminology, and reporting. Despite CT adding value in most of the identified studies, that value was relative to each individual situation and its extent could not be assessed with certainty. However, the usefulness of CT seems to depend on multiple factors including the research topic and feasibility/appropriateness of a primary database search. Our results support the use of multiple citation indexes in parallel and the conduct of several CT iterations but discourage from stand-alone CT in systematic literature searching. Indirect CT methods show great promise but require further research on refinement to be feasible. Based on our results and conclusions, we plan a Delphi study to develop consensus recommendations for the use and reporting of CT.^11^

## REQUIRED section

### What is already known

Citation tracking (CT) is an umbrella term and can be sub-categorized into direct and indirect CT methods. The added value of any form of CT to systematic literature searching is not clear.

### What is new

The benefit of CT likely depends on multiple factors that could not be assessed with certainty by synthesizing the collected evidence. Ample methodological heterogeneity among CT studies exemplifies the strong need for approved guidelines for conduct and reporting of CT.

### Potential impact for Research Synthesis Methods readers outside the authors’ field

For systematic reviews and other study designs aiming at a comprehensive retrieval of available evidence, the use of forward and backward CT on eligible articles should be considered as supplementary search methods. For non-systematic literature retrieval, any form of CT as a stand-alone search approach that is based on articles that are already known can be a valuable strategy.

## Supporting information

Supplementary material (data set)

## Data Availability

All data produced in the present work are contained in the manuscript

## Supporting information

- Supporting information 1: Database-specific search strategies
- Supporting information 2: Raw data set (separate file)

## Supporting information 1: Database-specific search strategies

### MEDLINE via Ovid (search date: October 26, 2020), 2,240 hits

(reference list or reference lists or ((reference OR references OR citation or citations or co-citation or co-citations) ADJ3 (search OR searches OR searching OR searched OR screen or screening or chain OR chains OR chaining OR check OR checking OR checked OR chased OR chasing OR tracking OR tracked OR harvesting OR tool or tools or backward or forward)) or ((cited OR citing OR cocited OR cociting OR co-cited OR co-citing) ADJ3 (references or reference)) or citation discovery tool or cocitation or co-citation or cocitations or co-citations or co-cited OR backward chaining or forward chaining or snowball sampling or snowballing or footnote chasing or berry picking or cross references or cross referencing or cross-references or cross-referencing or citation activity or citation activities or citation analysis or citation analyses or citation network or citation networks or citation relationship or citation relationships).ti OR (((((strategy or strategies OR method* OR literature OR evidence OR additional OR complementary OR supplementary) ADJ3 (find OR finding OR search* OR retriev*)) or (database ADJ2 combin*)).ti) AND ((search OR searches OR searching OR searched).ab))

### CINAHL (search date: October 26, 2020), 923 hits

(TI “reference list” OR TI “reference lists” OR ((TI reference OR TI references OR TI citation OR TI citations OR TI co-citation OR TI co-citations) N3 (TI search OR TI searches OR TI searching OR TI searched OR TI screen OR TI screening OR TI chain OR TI chains OR TI chaining OR TI check OR TI checking OR TI checked OR TI chased OR TI chasing OR TI tracking OR TI tracked OR TI harvesting OR TI tool OR TI tools OR TI backward OR TI forward)) OR ((TI cited OR TI citing OR TI cocited OR TI cociting OR TI co-cited OR TI co-citing) N3 (TI references OR TI reference)) OR TI “citation discovery tool” OR TI cocitation OR TI co-citation OR TI cocitations OR TI co-citations OR TI co-cited OR TI “backward chaining” OR TI “forward chaining” OR TI “snowball sampling” OR TI snowballing OR TI “footnote chasing” OR TI “berry picking” OR TI “cross references” OR TI “cross referencing” OR TI cross-references OR TI cross-referencing OR TI “citation activity” OR TI “citation activities” OR TI “citation analysis” OR TI “citation analyses” OR TI “citation network” OR TI “citation networks” OR TI “citation relationship” OR TI “citation relationships”) OR (((((TI strategy OR TI strategies OR TI method* OR TI literature OR TI evidence OR TI additional OR TI complementary OR TI supplementary) N3 (TI find OR TI finding OR TI search* OR TI retriev*)) OR (TI database N2 TI combin*))) AND ((AB search OR AB searches OR AB searching OR AB searched)))

### LLISFT (Library Literature & Information Science Full Text (search date: October 26, 2020), 973 hits

(TI “reference list” OR TI “reference lists” OR ((TI reference OR TI references OR TI citation OR TI citations OR TI co-citation OR TI co-citations) N3 (TI search OR TI searches OR TI searching OR TI searched OR TI screen OR TI screening OR TI chain OR TI chains OR TI chaining OR TI check OR TI checking OR TI checked OR TI chased OR TI chasing OR TI tracking OR TI tracked OR TI harvesting OR TI tool OR TI tools OR TI backward OR TI forward)) OR ((TI cited OR TI citing OR TI cocited OR TI cociting OR TI co-cited OR TI co-citing) N3 (TI references OR TI reference)) OR TI “citation discovery tool” OR TI cocitation OR TI co-citation OR TI cocitations OR TI co-citations OR TI co-cited OR TI “backward chaining” OR TI “forward chaining” OR TI “snowball sampling” OR TI snowballing OR TI “footnote chasing” OR TI “berry picking” OR TI “cross references” OR TI “cross referencing” OR TI cross-references OR TI cross-referencing OR TI “citation activity” OR TI “citation activities” OR TI “citation analysis” OR TI “citation analyses” OR TI “citation network” OR TI “citation networks” OR TI “citation relationship” OR TI “citation relationships”) OR (((((TI strategy OR TI strategies OR TI method* OR TI literature OR TI evidence OR TI additional OR TI complementary OR TI supplementary) N3 (TI find OR TI finding OR TI search* OR TI retriev*)) OR (TI database N2 TI combin*))) AND ((AB search OR AB searches OR AB searching OR AB searched)))

### LISTA via EBSCOhost (search date: October 26, 2020), 2,208 hits

(TI “reference list” OR TI “reference lists” OR ((TI reference OR TI references OR TI citation OR TI citations OR TI co-citation OR TI co-citations) N3 (TI search OR TI searches OR TI searching OR TI searched OR TI screen OR TI screening OR TI chain OR TI chains OR TI chaining OR TI check OR TI checking OR TI checked OR TI chased OR TI chasing OR TI tracking OR TI tracked OR TI harvesting OR TI tool OR TI tools OR TI backward OR TI forward)) OR ((TI cited OR TI citing OR TI cocited OR TI cociting OR TI co-cited OR TI co-citing) N3 (TI references OR TI reference)) OR TI “citation discovery tool” OR TI cocitation OR TI co-citation OR TI cocitations OR TI co-citations OR TI co-cited OR TI “backward chaining” OR TI “forward chaining” OR TI “snowball sampling” OR TI snowballing OR TI “footnote chasing” OR TI “berry picking” OR TI “cross references” OR TI “cross referencing” OR TI cross-references OR TI cross-referencing OR TI “citation activity” OR TI “citation activities” OR TI “citation analysis” OR TI “citation analyses” OR TI “citation network” OR TI “citation networks” OR TI “citation relationship” OR TI “citation relationships”) OR (((((TI strategy OR TI strategies OR TI method* OR TI literature OR TI evidence OR TI additional OR TI complementary OR TI supplementary) N3 (TI find OR TI finding OR TI search* OR TI retriev*)) OR (TI database N2 TI combin*))) AND ((AB search OR AB searches OR AB searching OR AB searched)))

### Web of Science Core Collection (search date: October 26, 2020) 5,754 hits

TI=((“reference list” OR “reference lists” OR ((reference OR references OR citation OR citations OR co-citation OR co-citations) NEAR/3 (search OR searches OR searching OR searched OR screen OR screening OR chain OR chains OR chaining OR check OR checking OR checked OR chased OR chasing OR tracking OR tracked OR harvesting OR tool OR tools OR backward OR forward)) OR ((cited OR citing OR cocited OR cociting OR co-cited OR co-citing) NEAR/3 (references OR reference)) OR “citation discovery tool” OR cocitation OR co-citation OR cocitations OR co-citations OR co-cited OR “backward chaining” OR “forward chaining” OR “snowball sampling” OR snowballing OR “footnote chasing” OR “berry picking” OR “cross references” OR “cross referencing” OR cross-references OR cross-referencing OR “citation activity” OR “citation activities” OR “citation analysis” OR “citation analyses” OR “citation network” OR “citation networks” OR “citation relationship” OR “citation relationships”)) OR (TI=((((strategy OR strategies OR method* OR literature OR evidence OR additional OR complementary OR supplementary) NEAR/3 (find OR finding OR search* OR retriev*)) OR (database NEAR/2 combin*))) AND (TS=(search OR searches OR searching OR searched)))

AND

(SU=(Life Sciences & Biomedicine OR Allergy OR Anatomy & Morphology OR Anesthesiology OR Biomedical Social Sciences OR Cardiovascular System & Cardiology ORCritical Care Medicine OR Dentistry, Oral Surgery & Medicine OR Dermatology OR Emergency Medicine OR Psychology OR Endocrinology & Metabolism OR Gastroenterology & Hepatology OR Food Science & Technology OR General & Internal Medicine OR Geriatrics & Gerontology OR Health Care Sciences & Services OR Hematology OR Immunology OR Infectious Diseases OR Information Science & Library Science OR Integrative & Complementary Medicine OR Legal Medicine OR Life Sciences Biomedicine Other Topics OR Mathematical & Computational Biology OR Medical Ethics OR Medical Informatics OR Neurosciences & Neurology OR Nursing OR Nutrition & Dietetics OR Obstetrics & Gynecology OR Oncology OR Ophthalmology OR Orthopedics OR Otorhinolaryngology OR Parasitology OR Pathology OR Pediatrics OR Pharmacology & Pharmacy OR Physiology OR Psychiatry OR Public, Environmental & Occupational Health OR Radiology, Nuclear Medicine & Medical Imaging OR Rehabilitation OR Research & Experimental Medicine OR Respiratory System OR Rheumatology OR Science & Technology Other Topics OR Sport Sciences OR Substance Abuse OR Surgery OR Toxicology OR Transplantation OR Tropical Medicine OR Urology & Nephrology OR Virology OR Behavioral Sciences OR Biochemistry & Molecular BiologyBiophysics OR Biotechnology & Applied Microbiology OR Cell Biology OR Genetics & Heredity OR Microbiology OR Reproductive Biology) OR WC=(Allergy OR Anatomy & Morphology OR Andrology OR Anesthesiology OR Audiology & Speech-Language Pathology OR Cardiac & Cardiovascular Systems OR Chemistry, Medicinal OR Clinical Neurology OR Critical Care Medicine OR Dentistry, Oral Surgery & Medicine OR Dermatology OR Emergency Medicine OR Endocrinology & Metabolism OR Engineering, Biomedical OR Ergonomics OR Food Science & Technology OR Gastroenterology & Hepatology OR Genetics & Heredity OR Geriatrics & Gerontology OR Gerontology OR Health Care Sciences & Services OR Health Policy & Services OR Hematology OR Immunology OR Infectious Diseases OR Information Science & Library Science OR Integrative & Complementary Medicine OR Mathematical & Computational Biology OR Medical Ethics OR Medical Informatics OR Medical Laboratory Technology OR Medicine, General & Internal OR Medicine, Legal OR Medicine, Research & Experimental OR Multidisciplinary Sciences OR Mycology OR Neuroimaging OR Neurosciences OR Nursing OR Nutrition & Dietetics OR Oncology OR Ophthalmology OR Orthopedics OR Otorhinolaryngology OR Parasitology OR Pathology OR Pediatrics OR Peripheral Vascular Disease OR Pharmacology & Pharmacy OR Physiology OR Primary Health Care OR Psychiatry OR Psychology OR Psychology, Multidisciplinary OR Psychology, Applied OR Psychology, Clinical OR Psychology, Experimental OR Psychology, Psychoanalysis OR Public, Environmental & Occupational Health OR Radiology, Nuclear Medicine & Medical Imaging OR Rehabilitation OR Respiratory System OR Rheumatology OR Social Sciences, Biomedical OR Sport Sciences OR Substance Abuse OR Surgery OR Toxicology OR Transplantation OR Tropical Medicine OR Urology & Nephrology OR Virology OR Behavioral Sciences OR Biochemical Research Methods OR Biochemistry & Molecular Biology OR Biophysics OR Biotechnology & Applied Microbiology OR Cell & Tissue Engineering OR Cell Biology OR Evolutionary Biology OR Microbiology OR Reproductive Biology))

## Supporting information 3: Raw data set

The file containing the raw data is separated from this text file.

## References

1. Sutton A, Clowes M, Preston L, Booth A. Meeting the review family: exploring review types and associated information retrieval requirements. Health Info Libr J. 2019;36(3):202–222. doi:10.1111/hir.12276

2. Lefebvre C, Glanville J, Briscoe S, et al. Searching for and selecting studies. In: Higgins JPT, Thomas J, eds. Cochrane Handbook for Systematic Reviews of Interventions: Version 6. 2nd ed. Wiley Online Library; 2019:67–108.

3. Ioannidis JPA. The Mass Production of Redundant, Misleading, and Conflicted Systematic Reviews and Meta-analyses. The Milbank Quarterly. 2016;94(3):485–514. doi:10.1111/1468-0009.12210

4. Abbas Z, Raza S, Ejaz K. Systematic Reviews and their role in Evidence - Informed Health Care. J Pak Med Assoc. 2008;58(10):561-567. Accessed December 16, 2019.

5. Gough D, Davies P, Jamtvedt G, et al. Evidence Synthesis International (ESI): Position Statement. Syst Rev. 2020;9:155. doi:10.1186/s13643-020-01415-5

6. Gusenbauer M, Haddaway NR. Which Academic Search Systems are Suitable for Systematic Reviews or Meta-Analyses? Evaluating Retrieval Qualities of Google Scholar, PubMed and 26 other Resources. Res Syn Meth. 2020;11:181–217. doi:10.1002/jrsm.1378

7. McGowan J, Sampson M. Systematic reviews need systematic searchers. J Med Libr Assoc. 2005;93(1):74–80.

8. Souza Leão L de, Eyal G. The rise of randomized controlled trials (RCTs) in international development in historical perspective. Theor Soc. 2019;48(3):383–418. doi:10.1007/s11186-019-09352-6

9. Cooper C, Booth A, Varley-Campbell J, Britten N, Garside R. Defining the process to literature searching in systematic reviews: a literature review of guidance and supporting studies. BMC Med Res Methodol. 2018;18:85. doi:10.1186/s12874-018-0545-3

10. Cooper C, Booth A, Britten N, Garside R. A comparison of results of empirical studies of supplementary search techniques and recommendations in review methodology handbooks: a methodological review. Syst Rev. 2017;6:234. doi:10.1186/s13643-017-0625-1

11. Hirt J, Nordhausen T, Appenzeller-Herzog C, Ewald H. Using citation tracking for systematic literature searching - study protocol for a scoping review of methodological studies and a Delphi study [version 3; peer review: 2 approved]. F1000Res. 2021;9:1386. doi:10.12688/f1000research.27337.3

12. Booth A. Unpacking your literature search toolbox: on search styles and tactics. Health Info Libr J. 2008;25(4):313–317. doi:10.1111/j.1471-1842.2008.00825.x

13. Booth A. Innovative approaches to systematic reviewing. In: Levay P, Craven J, eds. Systematic Searching: Practical ideas for improving results. Facet Publishing; 2019:25–50.

14. Hu X, Rousseau R, Chen J. On the definition of forward and backward citation generations. Journal of Informetrics. 2011;5(1):27–36. doi:10.1016/j.joi.2010.07.004

15. Briscoe S, Bethel A, Rogers M. Conduct and reporting of citation searching in Cochrane systematic reviews: a cross-sectional study. Research Synthesis Methods. 2020;11:169–180. doi:10.1002/jrsm.1355

16. Arksey H, O’Malley L. Scoping studies: towards a methodological framework. Int J Soc. 2005;8(1):19–32. doi:10.1080/1364557032000119616

17. Munn Z, Peters MDJ, Stern C, Tufanaru C, McArthur A, Aromataris E. Systematic review or scoping review? Guidance for authors when choosing between a systematic or scoping review approach. BMC Med Res Methodol. 2018;18(1):143. doi:10.1186/s12874-018-0611-x

18. Tricco AC, Lillie E, Zarin W, et al. PRISMA Extension for Scoping Reviews (PRISMA-ScR): Checklist and Explanation. Ann Intern Med. 2018;169(7):467–473. doi:10.7326/M18-0850

19. McGowan J, Sampson M, Salzwedel DM, Cogo E, Foerster V, Lefebvre C. PRESS peer review of electronic search strategies: 2015 guideline statement. J Clin Epidemiol. 2016;75:40–46. doi:10.1016/j.jclinepi.2016.01.021

20. Glanville J. Text mining for information specialists. In: Levay P, Craven J, eds. Systematic Searching: Practical ideas for improving results. Facet Publishing; 2019:147–170.

21. Clark JM, Sanders S, Carter M, et al. Improving the translation of search strategies using the Polyglot Search Translator: a randomized controlled trial. J Med Libr Assoc. 2020;108(2). doi:10.5195/jmla.2020.834

22. Bramer WM, Giustini D, Jonge GB de, Holland L, Bekhuis T. De-duplication of database search results for systematic reviews in EndNote. J Med Libr Assoc. 2016;104(3):240–243. doi:10.3163/1536-5050.104.3.014

23. Swiss Academic Software. Citavi. Accessed May 10, 2022. https://www.citavi.com/en

24. Ouzzani M, Hammady H, Fedorowicz Z, Elmagarmid A. Rayyan-a web and mobile app for systematic reviews. Syst Rev. 2016;5:210. doi:10.1186/s13643-016-0384-4

25. SWITCH. SWITCHdrive. Accessed May 15, 2022. https://www.switch.ch/drive/

26. Avenell A, Handoll HH, Grant AM. Lessons for search strategies from a systematic review, in The Cochrane Library, of nutritional supplementation trials in patients after hip fracture. Am J Clin Nutr. 2001;73(3):505–510. doi:10.1093/ajcn/73.3.505

27. Bae JM, Kim EH. Citation Discovery Tools for Conducting Adaptive Meta-analyses to Update Systematic Reviews. J Prev Med Public Health. 2016;49(2):129–133. doi:10.3961/jpmph.15.074

28. Bascur JP, Verberne S, van Eck NJ, Waltman L. Browsing Citation Clusters for Academic Literature Search: A Simulation Study with Systematic Reviews. BIR 2020 Workshop on Bibliometric-enhanced Information Retrieval. 2020:53–65.

29. Belter CW. Citation analysis as a literature search method for systematic reviews. J ASSOC INF SCI TECH. 2016;67(11):2766–2777. doi:10.1002/asi.23605

30. Belter CW. A relevance ranking method for citation-based search results. Scientometrics. 2017;112(2):731–746. doi:10.1007/s11192-017-2406-y

31. Boulos L, Ogilvie R, Hayden JA. Search methods for prognostic factor systematic reviews: a methodologic investigation. JMLA. 2021;109(1):23–32. doi:10.5195/jmla.2021.939

32. Chapman AL, Morgan LC, Gartlehner G. Semi-automating the manual literature search for systematic reviews increases efficiency. Health Info Libr J. 2010;27(1):22–27. doi:10.1111/j.1471-1842.2009.00865.x

33. Choong MK, Galgani F, Dunn AG, Tsafnat G. Automatic evidence retrieval for systematic reviews. J Med Internet Res. 2014;16(10):e223. doi:10.2196/jmir.3369

34. Cooper C, Lovell R, Husk K, Booth A, Garside R. Supplementary search methods were more effective and offered better value than bibliographic database searching: A case study from public health and environmental enhancement. Res Syn Meth. 2018;9(2):195–223. doi:10.1002/jrsm.1286

35. Goossen K, Hess S, Lunny C, Pieper D. Database combinations to retrieve systematic reviews in overviews of reviews: a methodological study. BMC Med Res Methodol. 2020;20:138. doi:10.1186/s12874-020-00983-3

36. Greenhalgh T, Peacock R. Effectiveness and efficiency of search methods in systematic reviews of complex evidence. BMJ. 2005;331(7524):1064–1065. doi:10.1136/bmj.38636.593461.68

37. Harari MB, Parola HR, Hartwell CJ, Riegelman A. Literature searches in systematic reviews and meta-analyses: A review, evaluation, and recommendations. J Vocat Behav. 2020;118:103377. doi:10.1016/j.jvb.2020.103377

38. Hinde S, Spackman E. Bidirectional citation searching to completion: an exploration of literature searching methods. Pharmacoeconomics. 2015;33(1):5–11. doi:10.1007/s40273-014-0205-3

39. Hirt J, Bergmann J, Karrer M. Overlaps of multiple database retrieval and citation tracking in dementia care research: a methodological study. J Med Libr Assoc. 2021;109(2):275–285. doi:10.5195/jmla.2021.1129

40. Janssens C. Updating systematic reviews and meta-analyses, the easy way. Accessed November 1, 2021. https://cecilejanssens.medium.com/updating-systematic-reviews-and-meta-analyses-the-easy-way-cbb2e23b48b9

41. Janssens AC, Gwinn M. Novel citation-based search method for scientific literature: application to meta-analyses. BMC Med Res Methodol. 2015;15:84. doi:10.1186/s12874-015-0077-z

42. Janssens A, Gwinn M, Brockman JE, Powell K, Goodman M. Novel citation-based search method for scientific literature: a validation study. BMC Med Res Methodol. 2020;20:25. doi:10.1186/s12874-020-0907-5

43. Kleijnen J, Knipschild P. The comprehensiveness of Medline and Embase computer searches. Searches for controlled trials of homoeopathy, ascorbic acid for common cold and ginkgo biloba for cerebral insufficiency and intermittent claudication. Pharm Weekbl Sci Ed. 1992;14(5):316–320. doi:10.1007/BF01977620

44. Kuper H, Nicholson A, Hemingway H. Searching for observational studies: what does citation tracking add to PubMed? A case study in depression and coronary heart disease. BMC Med Res Methodol. 2006;6:4. doi:10.1186/1471-2288-6-4

45. Lampert U, Waffenschmidt S. Benefit of additional search techniques to support literature searches in systematic reviews: the “forward citation searching” and “similar articles” functions. 15th EAHIL 2016 Conference. 2016.

46. Larsen B. Exploiting citation overlaps for Information Retrieval: Generating a boomerang effect from the network of scientific papers. Scientometrics. 2002;54(2):155–178. doi:10.1023/A:1016011326300

47. Lemeshow AR, Blum RE, Berlin JA, Stoto MA, Colditz GA. Searching one or two databases was insufficient for meta-analysis of observational studies. J Clin Epidemiol. 2005;58(9):867–873. doi:10.1016/j.jclinepi.2005.03.004

48. Levay P, Ainsworth N, Kettle R, Morgan A. Identifying evidence for public health guidance: a comparison of citation searching with Web of Science and Google Scholar. Res Syn Meth. 2016;7(1):34–45. doi:10.1002/jrsm.1158

49. Linder SK, Kamath GR, Pratt GF, Saraykar SS, Volk RJ. Citation searches are more sensitive than keyword searches to identify studies using specific measurement instruments. J Clin Epidemiol. 2015;68(4):412–417. doi:10.1016/j.jclinepi.2014.10.008

50. Lowe J, Peters J, Shields B, Cooper C. Methods to update systematic literature searches: full update searching vs. forward citation chasing: A case study from a systematic review of diagnostic test accuracy. InterTASC ISSG Workshop. 2014. https://medicine.exeter.ac.uk/media/universityofexeter/medicalschool/research/pentag/documents/J_Lowe_Poster_ISSG_Final.pdf

51. Matthews EJ, Edwards AG, Barker J, et al. Efficient literature searching in diffuse topics: lessons from a systematic review of research on communicating risk to patients in primary care. Health Libr Rev. 1999;16(2):112–120. doi:10.1046/j.1365-2532.1999.00219.x

52. McCain KW. Descriptor and citation retrieval in the medical behavioral sciences literature: retrieval overlaps and novelty distribution. Journal of the American Society for Information Science. 1989;40(2):110–114. doi:10.1002/(SICI)1097-4571(198903)40:2<110:AID-ASI5>3.0.CO;2-T

53. McManus RJ, Wilson S, Delaney BC, et al. Review of the usefulness of contacting other experts when conducting a literature search for systematic reviews. BMJ. 1998;317(7172):1562–1563. doi:10.1136/bmj.317.7172.1562

54. McNally R, Alborz A. Developing methods for systematic reviewing in health services delivery and organization: an example from a review of access to health care for people with learning disabilities. Part 1. Identifying the literature. Health Info Libr J. 2004;21(3):182–192. doi:10.1111/j.1471-1842.2004.00512.x

55. Murphy LS, Reinsch S, Najm WI, et al. Spinal palpation: the challenges of information retrieval using available databases. Physiol Ther J Manip Physiol Ther. 2003;26(6):374–382. doi:10.1016/S0161-4754(03)00076-9

56. Nussbaumer-Streit B, Klerings I, Wagner G, et al. Abbreviated literature searches were viable alternatives to comprehensive searches: a meta-epidemiological study. J Clin Epidemiol. 2018;102:1–11. doi:10.1016/j.jclinepi.2018.05.022

57. Ogilvie D, Hamilton V, Egan M, Petticrew M. Systematic reviews of health effects of social interventions: 1. Finding the evidence: how far should you go? J Epidemiol Community Health. 2005;59(9):804–808. doi:10.1136/jech.2005.034181

58. Pao ML. Comparing retrievals by keywords and citations. Proceedings - National Online Meeting. 1986:341–346.

59. Pao ML. Perusing the literature via citation links. Comput Biomed Res. 1993;26(2):143–156. doi:10.1006/cbmr.1993.1009

60. Pao ML. Term and citation retrieval: A field study. Inf Process Manag. 1993;29(1):95–112.

61. Pao ML, Fu TTW. Title retrieved from MEDLINE and from citation relations. Proceedings of the 48th ASIS Annual Meeting. 1985;22:120–123.

62. Pao ML, Worthen DB. Retrieval effectiveness by semantic and citation searching. J Am Soc Inf Sci. 1989;40(4):226–235. doi:10.1002/(SICI)1097-4571(198907)40:4<226:AID-ASI2>3.0.CO;2-6

63. Papaioannou D, Sutton A, Carroll C, Booth A, Wong R. Literature searching for social science systematic reviews: consideration of a range of search techniques. Health Info Libr J. 2010;27(2):114–122. doi:10.1111/j.1471-1842.2009.00863.x

64. Preston L, Carroll C, Gardois P, Paisley S, Kaltenthaler E. Improving search efficiency for systematic reviews of diagnostic test accuracy: an exploratory study to assess the viability of limiting to MEDLINE, EMBASE and reference checking. Syst Rev. 2015;4:82. doi:10.1186/s13643-015-0074-7

65. Rogers M, Bethel A, Briscoe S. Resources for forwards citation searching for implementation studies in dementia care: A case study comparing Web of Science and Scopus. Res Syn Meth. 2020;11(3):379–386. doi:10.1002/jrsm.1400

66. Sarol MJ, Liu L, Schneider J. Testing a citation and text-based framework for retrieving publications for literature reviews. BIR 2018 Workshop on Bibliometric-enhanced Information Retrieval. 2018:22–33.

67. Savoie I, Helmer D, Green CJ, Kazanjian A. Beyond MEDLINE: Reducing bias through extended systematic review search. Int J of Technology Assessment in Health Care. 2003;19(1):168–178. doi:10.1017/s0266462303000163

68. Stevinson C, Lawlor DA. Searching multiple databases for systematic reviews: added value or diminishing returns? Complement Ther Med. 2004;12(4):228–232. doi:10.1016/j.ctim.2004.09.003

69. Turki H. Automatic selection of references for the creation of a biomedical literature review using citation mapping. 3rd International Conference on Engineering Sciences for Biology and Medecine. 2017.

70. van Loo J. Medical and psychological effects of unemployment: a ‘grey’ literature search. Health Libr Rev. 1985;2(2):55–62. doi:10.1046/j.1365-2532.1985.220055.x

71. Westphal A, Kriston L, Holzel LP, Harter M, Wolff A von. Efficiency and contribution of strategies for finding randomized controlled trials: a case study from a systematic review on therapeutic interventions of chronic depression. J Public Health Res. 2014;3(2):177. doi:10.4081/jphr.2014.177

72. Wright K, Golder S, Rodriguez-Lopez R. Citation searching: a systematic review case study of multiple risk behaviour interventions. BMC Med Res Methodol. 2014;14:73. doi:10.1186/1471-2288-14-73

73. Janssens AC, Gwinn M. Erratum to: Novel citation-based search method for scientific literature: application to meta-analyses. BMC Med Res Methodol. 2015;15:97. doi:10.1186/s12874-015-0093-z

74. Cooper C. Improving Literature Searching in Systematic Reviews: The Application of Tailored Literature Searching Compared to ‘The Conventional Approach’. [Doctoral Thesis]: University of Exeter; 2017.

75. Jordan JL. Additional Search Strategies May Not Be Necessary for a Rapid Systematic Review. Evid Based Libr Inf Pract. 2015;10(2):150–152.

76. Horsley T, Dingwall O, Sampson M. Checking reference lists to find additional studies for systematic reviews. Cochrane Database Syst Rev. 2011;(8):MR000026. doi:10.1002/14651858.MR000026.pub2

77. Bramer WM. Reference checking for systematic reviews using Endnote. J Med Libr Assoc. 2018;106(4):542–546. doi:10.5195/jmla.2018.489

78. Hausner E, Cooper C, Harboe I, Halfpenny N, Waffenschmidt S, Wong R. Value of using different search approaches. Accessed October 29, 2021. https://sites.google.com/york.ac.uk/sureinfo/home/value-of-using-different-search-approaches

79. Haddaway NR, Grainger MJ, Gray CT. citationchaser: a tool for transparent and efficient forward and backward citation chasing in systematic searching. Res Syn Meth. 2022. doi:10.1002/jrsm.1563

80. Smalheiser NR, Schneider J, Torvik VI, Fragnito DP, Tirk EE. The Citation Cloud of a biomedical article: a free, public, web-based tool enabling citation analysis. JMLA. 2022;110(1):103–108. doi:10.5195/jmla.2022.1117

81. Skolarus TA, Lehmann T, Tabak RG, Harris J, Lecy J, Sales AE. Assessing citation networks for dissemination and implementation research frameworks. Implement Sci. 2017;12:97. doi:10.1186/s13012-017-0628-2

82. Li J, Burnham JF, Lemley T, Britton RM. Citation analysis: Comparison of Web of Science®, Scopus™, SciFinder®, and Google Scholar. J Med Libr Assoc. 2010;7(3):196–217. doi:10.1080/15424065.2010.505518

83. Boyack KW, Klavans R. Co-citation analysis, bibliographic coupling, and direct citation: Which citation approach represents the research front most accurately? J Am Soc Inf Sci. 2010;61(12):2389–2404. doi:10.1002/asi.21419

84. Justesen T, Freyberg J, Schultz ANØ. Database Selection and Data Gathering Methods in Systematic Reviews on Qualitative Research Regarding Diabetes Mellitus - an Explorative Study: Preprint. 2020.

85. Justesen T, Freyberg J, Schultz ANØ. Database selection and data gathering methods in systematic reviews of qualitative research regarding diabetes mellitus - an explorative study. BMC Med Res Methodol. 2021;21:94. doi:10.1186/s12874-021-01281-2

86. Ruppen W, Derry S, McQuay HJ, Moore RA. Infection rates associated with epidural indwelling catheters for seven days or longer: systematic review and meta-analysis. BMC Palliat Care. 2007;6:3. doi:10.1186/1472-684X-6-3

87. Cribbin T. Citation Chain Aggregation: An Interaction Model to Support Citation Cycling. Proceedings of the 20th ACM International Conference on Information and Knowledge Management. 2011. doi:10.1145/2063576.2063913

88. Cribbin T. Augmenting Citation Chain Aggregation with Article Maps. CEUR Workshop Proceedings. 2014;1311. https://www.researchgate.net/publication/269696064_Augmenting_Citation_Chain_Aggregation_with_Article_Maps

89. Sampson M, Daniel R, Cogo E, Dingwall O. Sources of evidence to support systematic reviews in librarianship. JMLA. 2008;96(1):66–69. doi:10.3163/1536-5050.96.1.66

90. Woodman J, Harden A, Thomas J, Brunton J, Kavanagh J, Stansfield C. Searching for systematic reviews of the effects of social and environmental interventions: a case study of children and obesity. JMLA. 2010;98(2):140–146. doi:10.3163/1536-5050.98.2.006

91. Wright JM, Cottrell DJ, Mir G. Searching for religion and mental health studies required health, social science, and grey literature databases. J Clin Epidemiol. 2014;67(7):800–810. doi:10.1016/j.jclinepi.2014.02.017

